# Novel Loci (*EIF4A2, ADIPOQ, TPRG1*) for Triglyceride / High-density Lipoprotein Cholesterol Ratio Longitudinal Change (ΔTHR) among Subjects without Type 2 Diabetes: Evidence from the Long Life Family Study (LLFS) and the Framingham Heart Study (FHS) Offspring Cohort (OS)

**DOI:** 10.1101/2024.06.18.24309120

**Authors:** Lihua Wang, Siyu Wang, Jason A. Anema, Vaha A. Moghaddam, Yanli Lu, Shiow Lin, E. Warwick Daw, Allison L. Kuipers, Iva Miljkovic, Michael Brent, Gary Patti, Bharat Thygarajan, Joseph M. Zmuda, Michael A. Province, Ping An

**Author notes:** Corresponding author information: Lihua Wang, PhD Washington University School of Medicine 4444 Forest Park Ave, St. Louis, MO 63108.

## Abstract

**Aims/hypothesis:** Triglyceride (TG)/High density lipoprotein cholesterol (HDL-C) ratio (THR) represents a single surrogate predictor of hyperinsulinemia or insulin resistance that is associated with premature aging processes, risk of diabetes and increased mortality. To identify novel genetic loci for THR change over time (ΔTHR), we conducted genome-wide association study (GWAS) and genome-wide linkage scan (GWLS) among subjects of European ancestry who had complete data from two exams collected about seven years apart from the Long Life Family Study (LLFS, n=1384), a study with familial clustering of exceptional longevity in the US and Denmark.

**Methods:** Subjects with diabetes or using medications for dyslipidemia were excluded from this analysis. ΔTHR was derived using growth curve modeling, and adjusted for age, sex, field centers, and principal components (PCs). GWAS was conducted using a linear mixed model accounted for familial relatedness. Our linkage scan was built on haplotype-based IBD estimation with 0.5 cM average spacing.

**Results:** Heritability of ΔTHR was moderate (46%). Our GWAS identified a significant locus at the *LPL* (*p*=1.58e-9) for ΔTHR; this gene locus has been reported before influencing baseline THR levels. Our GWLS found evidence for a significant linkage with a logarithm of the odds (LODs) exceeding 3 on *3q28* (LODs=4.1). Using a subset of 25 linkage enriched families (pedigree-specific LODs>0.1), we assessed sequence elements under *3q28* and identified two novel variants (*EIF4A2/ADIPOQ*-rs114108468, *p*=5e-6, MAF=1.8%; *TPRG1*-rs16864075, *p*=3e-6, MAF=8%; accounted for ∼28% and ∼29% of the linkage, respectively, and 57% jointly). While the former variant was associated with *EIF4A2* (*p*=7e-5) / *ADIPOQ* (*p*=3.49e-2) RNA transcriptional levels, the latter variant was not associated with *TPRG1* (*p*=0.23) RNA transcriptional levels. Replication in FHS OS observed modest effect of these loci on ΔTHR. Of 188 metabolites from 13 compound classes assayed in LLFS, we observed multiple metabolites (e.g., DG.38.5, PE.36.4, TG.58.3) that were significantly associated with the variants (*p*<3e-4).

**Conclusions:** Our linkage-guided sequence analysis approach permitted our discovery of two novel gene variants *EIF4A2/ADIPOQ*-rs114108468 and *TPRG1*-rs16864075 on *3q28* for ΔTHR among subjects without diabetes selected for exceptional survival and healthy aging.

## Introduction

THR is an accurate surrogate marker of insulin resistance (IR), where a higher THR indicates a poor response of cells in muscles, fat, and liver to insulin [1]. IR is a well-known underlying pathogenesis of type 2 diabetes (T2D). T2D affects over half a billion people globally with its complications influencing cardiovascular system, kidney, and neurological organs [2]. In the US, the total cost of diabetes in 2022 is $412.9 billion including $306.6 billion of direct medical costs and $106.3 billion of indirect costs [3]. To prevent diabetes and reduce the cost, the genetic determinants regulating THR might provide novel insights for understanding the molecular mechanisms of the T2D development, and nominate potential diagnostic markers, as well as therapeutic targets. The largest genome-wide association study (GWAS) of THR to date has been conducted and identified 369 independent single nucleotide polymorphisms (SNPs) using a sample of 402,398 unrelated Europeans [4]. However, this GWAS analysis is confined to examining cross-sectional THR at one time point in unrelated samples, lacking the statistical power to unveil genetic variants contributing to the longitudinal change of THR (hereafter referred to as ΔTHR), especially in information-rich family data. The genetic regulation of ΔTHR in family-based cohorts remains elusive.

In order to pinpoint the genetic determinants of ΔTHR in family-based cohort, we utilized the NIA Long Life Family Study (LLFS) in the discovery phase. LLFS, a multicenter family-based cohort, measured THR in two visits spaced 5-7 years apart [5]. Two orthogonal analyses including genome-wide association study (GWAS; adjusting familial relatedness) and genome-wide linkage scan (GWLS; modeling familial relatedness) were employed to maximize our statistical power. The functional molecules (RNA transcripts and lipids) regulated by the identified genetic signals were also examined. FHS OS was used to replicate the variants identified in the discovery phase.

## Methods

### Cohort demographics

We performed initial discovery GWAS and GWLS utilizing participants from the LLFS. LLFS is a multinational, multicenter, and multigenerational family-based longitudinal study that recruited exceptional long-lived pedigrees. LLFS, predominantly of European descent, constitutes a total of four field centers including Boston University Medical Center in Boston (MA), Columbia College of Physicians and Surgeons in New York (NY), the University of Pittsburgh in Pittsburgh (PA), and the University of Southern Denmark field center. The Visit 1 in-person collected various anthropometric, blood pressure, physical performance, pulmonary function, as well as different blood tests at 2006 to 2009 in 4953 individuals from 539 families, whereas the Visit 2 repeated all Visit 1 protocols at 2014 to 2017 with additional carotid ultrasonography measures [5]. To estimate the random coefficient of individualized longitudinal change for the ratio of TG to HDL-C [6], we excluded subjects with diabetes or those taking medications for dyslipidemia at either Visit 1 or Visit 2, and used 1384 non-diabetic participants with TG and HDL-C assayed at both Visit 1 and Visit 2 (Table 1).

**Table 1.** Demorgraphic Information of samples by field centers.

| **Table 1. Demographic Information of samples by field centers** |  |  |  |  |  |
| --- | --- | --- | --- | --- | --- |
|  | **ALL** | **Boston** | **Columbia** | **Denmark** | **Pittsburgh** |
| **N** | 1384 | 358 | 199 | 545 | 282 |
| **Female (%)** | 821 (59.3) | 220 (61.5) | 120 (60.3) | 299 (54.9) | 182 (64.5) |
| **Age (mean ± SE)** | 61.9 ± 0.3 | 60.8 ± 0.6 | 63.6 ± 0.9 | 61.7 ± 0.4 | 62.6 ± 0.8 |
| **TG (mean ± SE)** | 101.23 ± 1.7 | 101 ± 3.6 | 101. ± 5.7 | 104. ± 2.7 | 95.4 ± 2.8 |
| **HDL-C (mean ± SE)** | 63.54 ± 0.5 | 63.3 ± 1.0 | 63.4 ± 1.3 | 64.3 ± 0.7 | 62.5 ± 1.0 |
| **ΔTHR (mean ± SE)** | 0.0 ± 2.7 | -0.1 ± 2.8 | -0.5 ± 2.8 | 0.4 ± 2.7 | -0.2 ± 2.7 |

Boston = Boston University Columbia = Columbia University Denmark = University of Southern Denmark Pittsburgh = University of Pittsburgh Age is age in years at baseline Visit 1; Values denote mean ± standard Error (SE) TG denotes Triglyceride assayed in mg/dl at baseline Visit 1 ΔTHR = longitudinal change of Triglyceride/HDL-Cholesterol estimated with values from Visit 1 and Visit 2

### FHS OS

To replicate our discovery findings in both GWAS and linkage region, we used the FHS OS data (https://www.framinghamheartstudy.org/). FHS OS is a longitudinal population-based study of Framingham (MA) residents [7, 8]. Our replication utilized the 859 samples from offspring cohorts who had TG and HDL-C assayed in both the examination 5 (average age: 54.63 ± 9.80) and 7 (average age: 61.54 ± 9.76). In FHS OS, TG levels were measured using automated enzymatic assay procedures, and HDL-C was quantified using Dextran sulfate-Mg2+ precipitation procedure [9]. Participants with diabetes or on medications for dyslipidemia at either examination 5 or 7 were excluded from our replication. As part of the National Heart, Lung and Blood Institute’s TOPMed phase I, WGS data from FHS OS were sequenced at >30× depth of coverage from the Broad Institute of the Massachusetts Institute of Technology and Harvard. The joint calling of all samples, along with quality control (QC) of variants and samples were performed by the TOPMed Informatics Resource Center at University of Michigan [10].

### Estimation of the THR and the ΔTHR

In LLFS, blood was drawn after at least eight hours of fast, and blood triglycerides (TG, mg/dl) and high-density lipoprotein-cholesterol (HDL-C, mg/dl) were quantified by the LLFS’s central laboratory at the University of Minnesota [11]. TG levels were determined using the glycerol-blanked method with the Roche Diagnostics system, while HDL-C was assessed using the third-generation HDL-C test also from Roche Diagnostics. Both Visit 1 and Visit 2 employed the same protocol for assaying TG, as well as HDL-C. The lab assays performed at two distinct time points spaced seven years apart for the same subset of LLFS samples that showed strong correlated values. Only samples who had TG and HDL-C levels from both Visit 1 and Visit 2 were included in our analyses. THR was calculated for LLFS Visit 1 and Visit 2 separately, and followed by logarithmic transformation to approximate a normal distribution (Figure 1A). The covariate effect of baseline age, field centers, sex, and 10 PCs (details see Estimation of principal components) were adjusted using a linear model in SAS 9.4. We performed a random coefficient model (RCM) using a linear mixed model in SAS 9.4 and estimated individualized intercepts and slope for the adjusted residuals. Under the assumption that the intercepts and slopes are multivariate and normally distributed with an unknown variance-covariance matrix, this approach simultaneously estimates inter-individual differences and intra-individual systematic changes over time and increases the precision of both intercept and slope estimates. Similarly, THR was calculated for non-diabetic participants who are not on dyslipidemia medication and had TG and HDL-C levels from both FHS OS examination 5 and 7, followed by logarithmic transformation, adjustment for baseline age and sex, and standardized ΔTHR estimation using a linear mixed model in SAS 9.4. Family relatedness was not considered in the estimation of ΔTHR here, and was adjusted in the association analyses.

**Figure 1.**
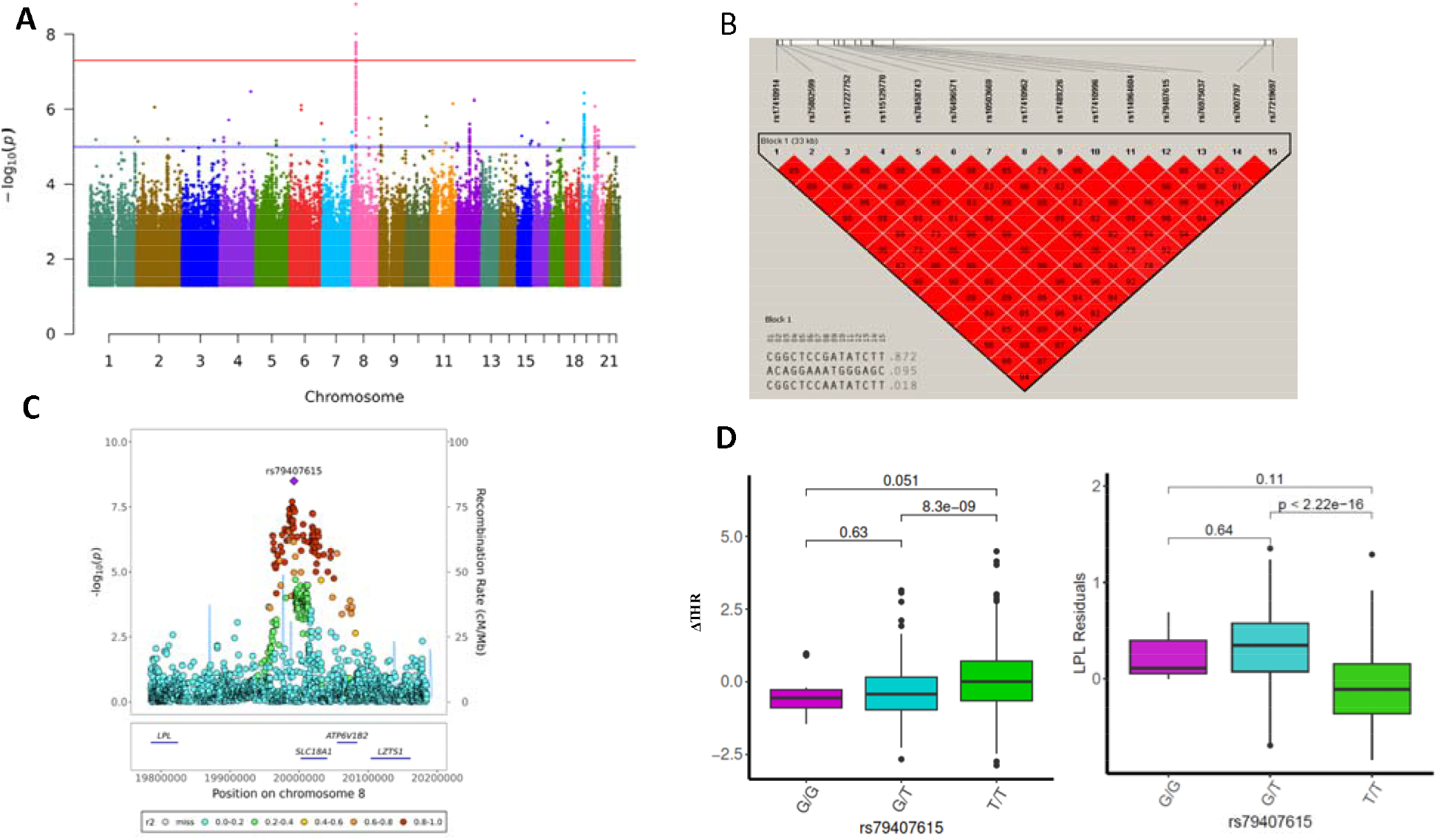
GWAS results of ΔTHR using whole genome sequenced autosome variants. **A)** The Manhantan plot for GWAS results of ΔTHR across 22 chromosomes. P values are two-sided raw P values estimated from a linear additive model. The y-axis depicts the negative log10-transformed P value. The x-axis is genomic coordinates by chromosome number. The blue-colored solid horizontal line denotes the suggestive threshold (*p*=1e-5). The red-colored solid horizontal line indicates the genome-wide significant cutoff at *p*=5e-8. **B)** The LD heatmap of 15 significant SNPs at chromosome 8 that reached genome-wide significance using Haploview. The value displayed is LD r2. **C)** The Locuszoom plot of ± 200 Kb of lead SNP rs79407615 at chromosome 8. The x-axis is the base pair position in the genome build GRCh38 at chromosome 8. The y-axis is the –log10 of the two-sided P value for each genetic variants. **D)** The box and whisker plot for the ΔTHR and the LPL residuals across three genotypes of rs79407615. The pairwise comparison P value is estimated using wilcox.test in R.

### GWAS genotyping in LLFS

We utilized the automated Autopure DNA extraction method (Qiagen Inc., Gaithersburg, MD) to extract DNA from white blood cells. The Center for Inherited Disease Research (CIDR) genotyped 2.5 million single-nucleotide polymorphisms (SNPs) using the Illumina Human Omni 2.5 v1 chips. An in-house QC of the genotypes was conducted by the Division of Statistical Genomics of Washington University. For SNP level QC, a total of 83,774 SNPs with call rate <98% were removed, and 3647 SNPs with a high Mendelian error rate detected by LOKI 2.4.5 [12] were dropped. In terms of subject level QC, 18 subjects with a call rate<97.5% were excluded. In addition, 153,363 data points had Mendelian errors and we set them to missing in the corresponding families.

### Whole Genome Sequencing (WGS) in LLFS

WGS in 150 bp paired-end reads was performed using Illumina sequencer by the McDonnell Genome Institute (MGI) at Washington University in Saint Louis. MGI also conducted reads alignment to Genome Reference Consortium Human Build 38 (GRCh38) with Burrows-Wheeler Aligner (BWA-MEM 0.7.15), marking duplicates with Picard 2.4.1, base quality score recalibration with GATK BaseRecalibrator 3.6, and lossless conversion to CRAM format with SAMtools 1.3.1. The variant calling was carried out at the Division of Statistical Genomics at Washington University in St. Louis[13]. The four steps of variant calling are: 1) CRAM files were converted to subject-level GVCF files with GATK HaplotypeCaller; 2) GVCF files were combined using GATK CombineGVCF; 3) Genotype of identified variants were called with GATK GenotypeGVCFs; 4) the diallelic SNPs were extracted using GATK SelectVariants. The additional QC procedures include exclusion of contaminated samples with FREEMIX >0.03, samples with <20x of haploid coverage, as well as samples with high Mendelian errors reported by LOKI 2.4.5 [12] and KING 2.3.1. The low quality variant site that had depth <20 or >300, Hardy-Weinberg equilibrium (HWE) *p*<1e-6, or heterozygosity>0.55 were removed. The sample swap was identified and corrected by comparing WGS against GWAS chip data. After QC, 1,720 participants with 32,892,186 autosome diallelic SNPs remained.

### RNA Sequencing (RNA-seq) for blood samples at LLFS visit 1

MGI extracted RNA from the PAXgene^TM^ Blood RNA tubes using the Qiagen PreAnalytiX PAXgene Blood miRNA Kit (Qiagen, Valencia, CA). The whole blood paired-end RNA sequencing library prep, quality control, alignment, and gene expression matrix were done by the Division of Computation & Data Sciences at Washington University[14]. The nf-core/rnaseq 3.14.0 was employed to obtain the quantification of the RNA sequencing data (https://nf-co.re/rnaseq/3.14.0). The major steps involve aligning the reads to GRCh38 with STAR, marking read duplicates with Picard MarkDuplicates, and transcript assembly and quantification with StringTie. We additionally removed the genes with fewer than four counts per million in at least 98.5% of samples, as well as the genes with more than 8% of intergenic overlap. After QC, 1810 samples with 16418 genes remained. The count of each gene were normalized using a variance stabilizing transformation (VST) from the fitted dispersion-mean relation in DESeq2.

### Measurement of blood lipid metabolites at LLFS visit 1

The untargeted metabolomics workflow for lipids was conducted at the Biomedical Mass Spectrometry Lab at Washington University [15]. In brief, lipid metabolites were extracted from plasma samples using a solid-phase extraction (SPE) plate with methyl tert-butyl ether/methanol. Subsequently, the lipid extract was dried using nitrogen flow. The *m/z* values of lipid metabolites were obtained via reversed-phase (RP) chromatography coupled to high-resolution mass spectrometry (HRMS) in positive mode. Lipid Annotator was employed for annotating the lipid iterative MS/MS data. The peak areas of the annotated peak list were quantified using Skyline [16] (version 20.1.0.155). Blank samples were introduced at the beginning and end of each batch to detect background peaks. To mitigate batch variation, a pooled QC sample served as an internal standard, and a random forest-based method was applied to correct peak areas [17]. After QC, 188 lipids from 13 compound classes remained.

### Estimation of principal components in LLFS

To examine the population structure in LLFS, we estimated principal components (PCs) using Eigenstrat [18]. To select the tag SNPs with good quality for estimating PCs, we removed the subsets with MAF<5%, HWE *p*<1e-6, as well as SNPs in some special regions (2q21, 2q21.1, HLA at chromosome 6, 8p23.1, 8p23, 17q21.31, and 17q21.311). Finally, a total of 116,867 tag SNPs present in both LLFS and HapMap data were chosen to estimate the PCs. Since LLFS is a family-based study and participants are not independent, we utilized a two-step approach to obtain the PCs. In the first step, 1522 unrelated subjects from LLFS and 361 founders from HapMap data including four different populations: Caucasians, Yorubans, Asians, and Toscani in Italia (TSI)-Caucasians were used to generate the top 20 PCs model. Then in the second step, PC estimators were expanded to all WGS genotyped members of LLFS using the Eigenstrat framework.

### GWAS using WGS Data in LLFS and replication in FHS OS

To identify the sequenced variants that are associated with ΔTHR, we conducted GWAS analyses and replication using a linear mixed model for additive dosage of the variants. We defined the major allele of each variant as the reference allele (REF). For instance, the dosage is 0 for genotype of two copies of reference allele (REF/REF), 1 for one copy of reference and one copy of alternative allele (REF/ALT), and 2 for two copies of alternative allele (ALT/ALT). To account for the familial relationship, the kinship matrix was estimated using “kinship” R package and included as a random effect in “lmekin” R package. The significant variants were identified with *p*<5e-8. The Manhattan plot and Quantile-quantile plots were generated using “qqman” package in R. The r^2^ measure of linkage disequilibrium (LD) was estimated in Haploview4.1.

### GWLS and fine mapping under the linkage peak in LLFS

To detect the genetic regions contributing to heritability of ΔTHR within families, we first selected up to five tightly linked SNPs within each of 0.5 centiMorgan (cM) interval across the autosome and built information rich haplotypes using ZAPLO [19]. From these haplotypes, we estimated multipoint identity-by-descent (IBD) using LOKI 2.4.5 [12] in intact pedigrees and performed GWLS via Sequential Oligogenic Linkage Analysis Routines (SOLAR). Any autosomal region with LODs above 3 was identified as significant linkage peak and selected for further analysis. As ΔTHR is a complex trait potentially influenced by multiple genes and their interactions, variation of ΔTHR among different families may be attributed to different genetic factors and regions. Consequently, due to genetic heterogeneity and varying penetrance across families, only selected LLFS families had pedigree-specific LODs exceeding 0.1 and contributed to the significant linkage peak. The LODs estimated using these selected families were defined as heterogeneity LODs (HLOD). To enhance the statistical power of our fine mapping and nominate potential driver SNPs, we focused on these selected families. To quantify the linkage peak explained by the top sequenced variants, we generated a dataset comprising ΔTHR and dosage with identical non-missing patterns for each of these variants. Next, we assessed the HLOD prior to SNP adjustment in SOLAR without any covariates, then obtained the HLOD after SNP adjustment by integrating the dosage of each SNP as a covariate in SOLAR. We subsequently calculated the percentage of linkage explained by the difference in HLOD before and after SNP adjustment divided by the HLOD before adjustment.

### Expression quantitative trait loci (eQTLs) analysis in LLFS

To map the sequenced variants to their regulated functional molecules, we first adjusted normalized values of their nearby genes by age, sex, field centers, white blood cell count, red blood cell count, platelets, monocyte, neutrophile, plates, percent of intergenic reads, and 10 principal components, and then tested the association of the sequenced variants with the adjusted residuals using a linear mixed model implemented in “kinship” and “lmekin” R package.

### Metabolite quantitative trait loci (mQTLs) analysis in LLFS

The variants influencing ΔTHR may operate through the regulation of blood levels of lipid metabolites. To assess the association of sequenced variants with lipid metabolite level in blood during the first visit, we log2-transformed the peak area of each lipid metabolite to approximate a normal distribution. Subsequently, we adjusted the transformed values for the effects of age, sex, field centers, and 10 principal components. The effect size of the sequenced variants on the adjusted residuals was determined using a linear mixed model implemented in “kinship” and “lmekin” R package.

## Results

### Characteristics of samples

As shown in Table 1, among 1384 samples, about 39.4% (545) are from the Denmark center, 25.8% (358) from the Boston center, 20.4% (282) from the Pittsburgh center, and 14.4% (199) from the Columbia center. Each of the four field centers included more than half the female participants: 64.54% (182) from the Pittsburgh center, 61.45% (220) from the Boston center, 60.3% (120) from the Columbia center, and 54.86% (299) from the Denmark center. The mean age of blood draw in the first visit of these 1384 individuals is 61.9 years old, ranging from 60.8 in the Boston center to 63.6 in the Columbia center. The average TG level of 1384 individuals is 101.23 mg/dl. Samples from the Pittsburgh center had relatively low levels of TG (95.4 mg/dl) than participants from the Boston center (101 mg/dl), Columbia center (101 mg/dl), and Denmark center (104 mg/dl). The mean HDL-C level of all samples is 63.54 mg/dl and varies slightly from 63.3 mg/dl in Boston center, 63.4 mg/dl in Columbia center, 64.3 mg/dl in Denmark center, and 62.5 mg/dl in Pittsburgh center. The mean ΔTHR level ranges from -0.5 in Columbia center to 0.4 in Denmark center. (Table 1).

### Discovery of *LPL* locus from GWAS

For the 1384 LLFS samples, we conducted a GWAS of ΔTHR using 7,944,836 sequenced SNPs with minor allele frequency (MAF) greater than 0.01. Fifteen common variants, located at 18,692 bp – 52,691 bp downstream of *LPL* at chromosome 8, reached the genome-wide significant threshold (*p*<5e-8; Figure 1A, 1C). Based on Haploview, these 15 variants are in LD with each other, all with pairwise r^2^>0.8 (Figure 1B). The SNP rs79407615 is the sentinel variant of the identified locus. We found that the G allele of rs79407615 is notably enriched in our samples (G=9.44%), which is almost four times the frequency of this allele in the NCBI Allele Frequency Aggregator (ALFA) European database (G=2.38%). The participants carrying G allele of rs79407615 had lower ΔTHR (β = -0.4123; P = 1.58×10^-9^; Figure 1D), indicating protective role of this SNP against the development of IR and T2D. In addition, the G allele of rs79407615 was also associated with lower THR in LLFS Visit 1 (β = -0.37; *p* = 4.65×10^-8^). To examine whether *LPL* is the functional molecule that mediates the effect of this SNP, we examined the association of rs79407615 with blood expression of *LPL*. We observed the G allele of rs79407615 was associated with significantly higher blood *LPL* expression (β=0.3233; *p*=3.004e-43; Figure 1D), highlighting *LPL* is the molecule that links rs79407615 to ΔTHR.

### Replication of *LPL* locus in FHS OS data

To replicate the association of *LPL* locus with ΔTHR, we estimated the random slope coefficient using 859 FHS OS non-diabetic participants in offspring cohorts who had TG and HDL-C assayed at both examination 5 and 7. We utilized the linear mixed model accounting for family relatedness. A total of 236 SNPs (50 independent signals with r^2^ ≤0.2) within the region of 15 significant variants (19,985,951 bp - 20,019,950 bp), and 314 SNPs (85 independent signals with r^2^ ≤0.2) within the region of *LPL* gene (19,901,717 bp -19,967,259 bp) were present in FHS OS WGS data. None of the SNPs in *LPL* locus passed the Bonferroni corrected threshold (0.05/50=1e-3 and 0.05/85=5.88e-4). Though not reaching nominal significance (*p*= 0.279), the SNP rs79407615 G allele had the consistent negative effect (β = - 0.02) on ΔTHR in FHS OS. In addition, a nearby rare SNP (G=0.79%) rs117956669, 2,454 bp downstream of our sentinel variant, shows nominal significance in association with lower ΔTHR (β=-0.184; *p*=5.9e-3). The variant rs117956669 is completely independent of rs79407615, with a negligible r^2^ value between them (r^2^= 0.00087). We also found a common intronic variant of *LPL*, rs112122343 (T=5.81%; β=0.079; *p*= 2.94e-3), is nominally significant for ΔTHR.

### Validation of previous reported loci influencing cross-sectional THR

Oliveri et al reported 369 independent SNPs for cross-sectional THR in a GWAS study using 402,398 Europeans [4]. About 96% (354/369) of these SNPs are available in LLFS WGS data. Among these 354 SNPs, we found 25 SNPs (7.06%) from 23 loci (*LPL*, *ANGPTL3*, *ANGPTL4*, and *TM6SF2*, as well as others), had *p*<0.05 and showed the same direction of effect for ΔTHR (Table 2). Notably, the SNP rs268, a missense variant of *LPL* (MAF=1.82%; β=0.58; *p*=1.11e-4) passed the Bonferroni corrected threshold (0.05/354=1.27e-4) for association with ΔTHR in LLFS, and its G allele led to a higher THR (β=0.29; *p*= 1.11e-136; Oliveri’s paper) and a higher ΔTHR (LLFS).

**Table 2.** Validation of reported loci for cross-sectional THR.

| rsID | chr | pos | Locus | role | CA | Non_CA | eaf | LLFS |  |  | Oliveri et al |  |  |
| --- | --- | --- | --- | --- | --- | --- | --- | --- | --- | --- | --- | --- | --- |
| | | | | | | | | $\beta$ | SE | P | $\beta$ | SE | P |
| rs10889333 | 1 | 62491359 | ANGPTL3 | intronic | A | G | 0.328 | -0.124 | 0.043 | 3.70E-03 | -0.066 | 0.003 | 3.37E-89 |
| rs13094241 | 3 | 196464022 | UBXN7-AS1 | intergenic | T | G | 0.262 | 0.096 | 0.046 | 3.61E-02 | 0.021 | 0.003 | 5.11E-09 |
| rs998584 | 6 | 43790159 | VEGFA | intergenic | A | C | 0.489 | 0.088 | 0.039 | 2.44E-02 | 0.062 | 0.003 | 8.55E-86 |
| rs12208357 | 6 | 160122116 | SLC22A1 | nonsynonymous_SNV | T | C | 0.068 | 0.168 | 0.078 | 3.04E-02 | 0.042 | 0.005 | 1.44E-11 |
| rs17145750 | 7 | 73612048 | MLXIPL | intronic | T | C | 0.167 | -0.110 | 0.054 | 4.28E-02 | -0.142 | 0.004 | 2.76E-241 |
| rs111914893 | 7 | 74587994 | GTF2IRD1 | intronic | T | C | 0.062 | 0.226 | 0.081 | 5.16E-03 | 0.041 | 0.006 | 1.51E-08 |
| rs10260148 | 7 | 130746210 | KLF14 | intergenic | T | C | 0.268 | 0.114 | 0.045 | 1.07E-02 | 0.043 | 0.003 | 9.46E-35 |
| rs62492368 | 7 | 150840547 | AOC1 | intergenic | A | G | 0.320 | 0.099 | 0.042 | 2.02E-02 | 0.022 | 0.003 | 8.89E-11 |
| rs7012814 | 8 | 9315848 | LOC157273 | intergenic | A | G | 0.468 | -0.113 | 0.040 | 5.35E-03 | -0.038 | 0.003 | 2.07E-33 |
| rs268 | 8 | 19956018 | LPL | nonsynonymous_SNV | G | A | 0.018 | 0.584 | 0.151 | 1.11E-04 | 0.292 | 0.010 | 1.11E-136 |
| rs73667496 | 8 | 20080341 | LPL,SLC18A1 | intergenic | T | C | 0.060 | -0.232 | 0.084 | 6.04E-03 | -0.099 | 0.006 | 6.59E-44 |
| rs7015766 | 8 | 20081538 | LPL,SLC18A1 | intergenic | T | C | 0.076 | -0.251 | 0.077 | 1.13E-03 | -0.263 | 0.005 | 8.39E-405 |
| rs6999569 | 8 | 125463528 | TRIB1 | intergenic | G | A | 0.488 | -0.106 | 0.041 | 9.19E-03 | -0.108 | 0.003 | 6.83E-258 |
| rs9919491 | 10 | 92995002 | EXOC6 | intronic | A | T | 0.275 | 0.103 | 0.047 | 2.98E-02 | 0.021 | 0.003 | 2.64E-09 |
| rs480823 | 11 | 116655013 | LINC02702 | ncRNA_intronic | C | T | 0.083 | 0.262 | 0.071 | 2.44E-04 | 0.206 | 0.005 | 2.09E-267 |
| rs117233107 | 12 | 4219355 | CCND2 | intergenic | A | G | 0.019 | -0.525 | 0.146 | 3.32E-04 | -0.088 | 0.011 | 9.84E-11 |
| rs72735627 | 15 | 40765309 | GCHFR | UTR3 | T | C | 0.120 | -0.149 | 0.062 | 1.70E-02 | -0.032 | 0.004 | 1.85E-09 |
| rs139974673 | 15 | 43735687 | MAP1A | downstream | C | T | 0.034 | 0.296 | 0.112 | 8.27E-03 | 0.197 | 0.008 | 2.51E-88 |
| rs72786786 | 16 | 56951602 | HERPUD1 | intergenic | A | G | 0.331 | -0.090 | 0.041 | 2.94E-02 | -0.151 | 0.003 | 4.12E-421 |
| rs7499892 | 16 | 56972678 | CETP | intronic | T | C | 0.169 | 0.162 | 0.053 | 2.33E-03 | 0.171 | 0.003 | 2.15E-381 |
| rs931992 | 17 | 39665182 | STARD3 | upstream;downstream | G | T | 0.344 | 0.096 | 0.042 | 2.28E-02 | 0.020 | 0.003 | 8.65E-10 |
| rs116843064 | 19 | 8364439 | ANGPTL4 | nonsynonymous_SNV | A | G | 0.024 | -0.441 | 0.131 | 7.67E-04 | -0.260 | 0.009 | 5.57E-116 |
| rs58542926 | 19 | 19268740 | TM6SF2 | nonsynonymous_SNV | T | C | 0.081 | -0.224 | 0.073 | 2.14E-03 | -0.108 | 0.005 | 7.28E-74 |
| rs185799410 | 20 | 58891038 | GNAS | intronic | T | G | 0.028 | 0.263 | 0.121 | 3.02E-02 | 0.056 | 0.008 | 2.57E-08 |
| rs2223041 | 21 | 15050282 | NRIP1 | intronic | T | C | 0.386 | 0.123 | 0.042 | 3.74E-03 | 0.018 | 0.003 | 5.00E-08 |

chr = chromosome number
pos = base pair positions
Locus = mapped gene locus
CA = the allele used to estimate the effect
Non\_CA = the other allele not used for estimation of the effect
eaf = the frequency of coded allele
$\beta$ = the effect estimate
SE = Standard Error
P = two-sided P value

### GWLS and fine mapping detected *EIF4A2/ADIPOQ*-rs114108468 and *TPRG1*-rs16864075 at 3q28

In LLFS, heritability of THR at two visits (40% in Visit 1 and 32% in Visit 2) was comparable to ΔTHR (46%). Our GWLS, conducted using SOLAR, identified a genomic region located at 3q28 with LODs score exceeding 3.0 (LODs=4.1; Figure 2A). At this linkage peak (184,989,149 bp – 192,579,505 bp), we selected 25 families encompassing 234 participants whose pedigree-specific LODs exceeded 0.1. Using these selected families, the HLOD score collectively reached 6.95 at 197 cM, significantly higher than the LODs score obtained from the analysis of 397 pedigrees. To pinpoint the genetic variants contributing to the linkage peak, we initially investigated the association of sequenced variants in the region with the ΔTHR of these 25 families using a linear mixed model, and followed by fine mapping. As shown in Figure 2B, we detected two variants, rs114108468 (133,231 bp upstream of *ADIPOQ;* 13,962 bp downstream of *EIF4A2*) and rs16864075 (an intronic variant of *TPRG1*), that had *p*<1.0e-4 for ΔTHR and a HLOD drop of ∼2. These two SNPs are independent and not in LD with each other (r^2^= 0.0003). The SNP rs114108468, a rare variant, had enriched G allele (G=1.5%) in our selected families than in ALFA European database (G=0.7%), and dosage of G allele correlated with a higher ΔTHR (β=1.965; *p*=5.68e-6; Figure 2C). The second SNP rs16864075 also showed higher G allele frequency (G=7.38%) in our samples than in ALFA European database (G=6.11%), and this G allele significantly increased the ΔTHR (β = 0.85; *p*=1.10e-6; Figure 2C). Further linkage analysis for fine mapping, conditioning on each of rs114108468 and rs16864075, revealed a HLOD drop of 1.94 (∼28% of linkage) by rs114108468 and 2.05 (∼29% of linkage) by rs16864075, supporting their significant contribution to the linkage peak. Subsequently, integration of blood transcriptomic data uncovered that the G allele of rs114108468 is associated with a significantly lower *ADIPOQ* level (β= -0.6134; *p*=3.49e-2; Figure 2C) and lower *EIF4A2* level (β=-0.1322; *p*=7.00e-5; Figure 2C) in the blood of LLFS participants, implicating *ADIPOQ* and *EIF4A2* in the regulation of ΔTHR. No notable association was found between rs16864075 and *TPRG1* (*p*=2.33e-1).

**Figure 2.**
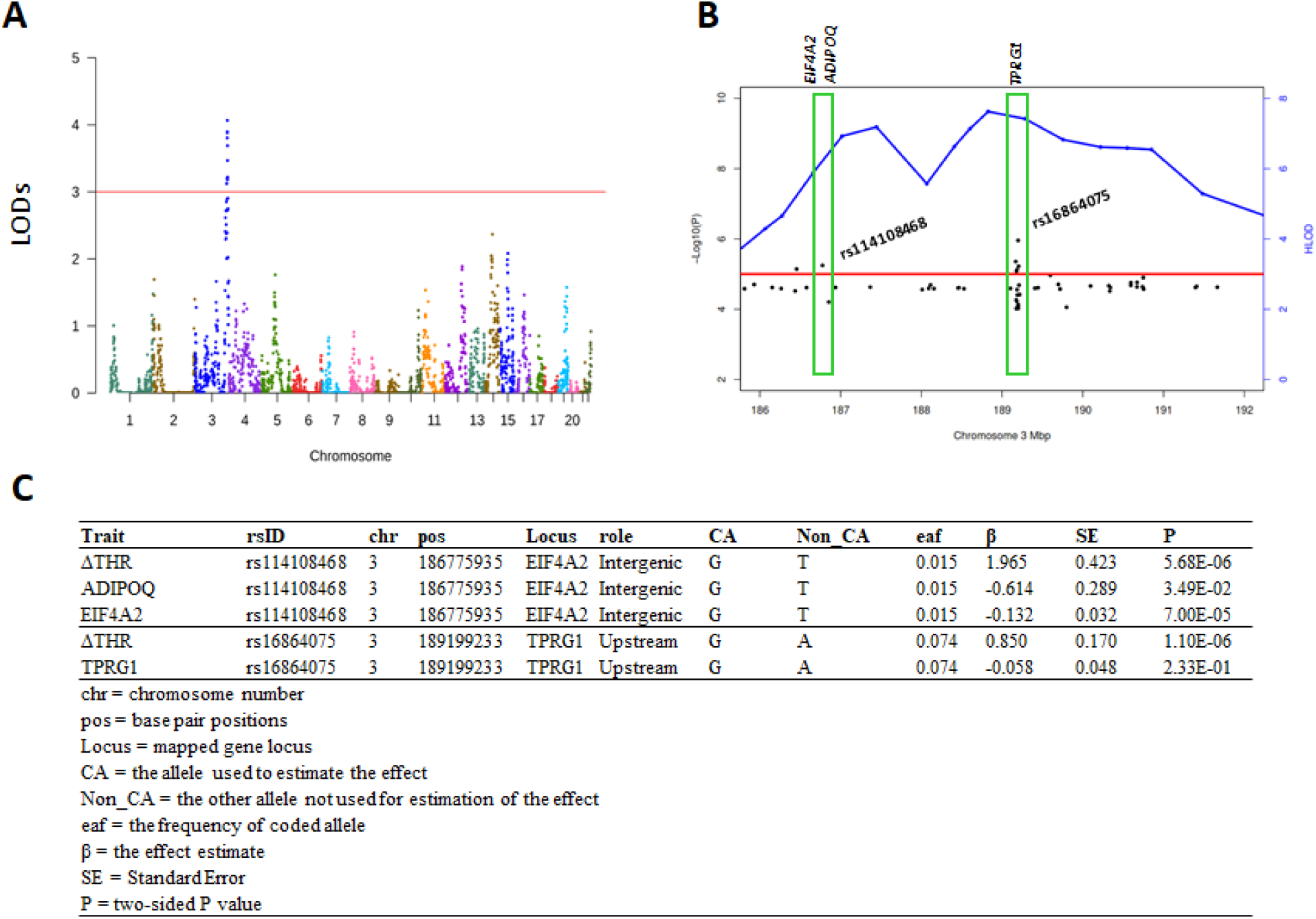
Genome-wide linkage analyses of the ΔTHR identified a linkage peak at chromosome 3. **A)** The plot of the LOD score across 22 autosomes. The Y-axis is the LOD score estimated with SOLAR. The X-axis is the genomic coordinates by chromosome number. **B)** The plot of negative log10 transformed *p* value (Y-axis at right side) and HLOD (Y-axis at left side) vs physical position in Mbp at chromosome 3. **C)** The association results of rs114108468 and rs16864075 with ΔTHR, *ADIPOQ*, *EIF4A2* and *TPRG1*.

### Replication of 3q28 locus in FHS OS data

The heritability of ΔTHR in FHS was 36%. To examine whether the association of 3q28 locus can be replicated in independent datasets, we analyzed the association of 3q28 locus with ΔTHR. Using 859 FHS OS non-diabetic offspring cohorts participants, we obtained the ΔTHR using a linear mixed model, and tested the association of sequenced variants that are ± 20 Kb of rs114108468 and rs16864075 with ΔTHR. Both of these SNPs at 3q28 locus did not reach nominal significance in FHS OS. However, rs189908673 (MAF=0.2%; β=0.277; *p*=1.27e-2; r^2^=0 with rs114108468) that is located 11,998 bp downstream of rs114108468, and rs113022387 (MAF=0.45%; β=0.1747; *p*=4.75e-2; r^2^=0.0001 with rs16864075) 835 bp upstream of rs16864075 had a weak association with ΔTHR.

### Multiple lipids associated with rs79407615, rs114108468, and rs16864075

TG is one of the components involved in the calculation of THR. To test whether the *LPL* locus and 3q28 locus harbors mQTLs of lipids, we examined the association of rs79407615, rs114108468 and rs16864075 with 188 lipids from blood at visit 1 using a linear mixed model adjusting for age, sex, field centers and 10 principal components. As shown in Table 3, 14 lipids from two compound classes (Diacylglycerol, and Triacylglycerol) for rs79407615, 22 lipids from four compound classes (Phosphatidylethanolamine, Diacylglycerol, Triacylglycerol, and Cholesteryl ester) for rs114108468, and nine lipids from three compound classes (Diacylglycerol, Triacylglycerol, and Cholesteryl ester) passed the Bonferroni corrected threshold (*p*<2.66e-4), supporting the involvement of *LPL* locus and 3q28 locus in the regulation of multiple lipids.

**Table 3.**
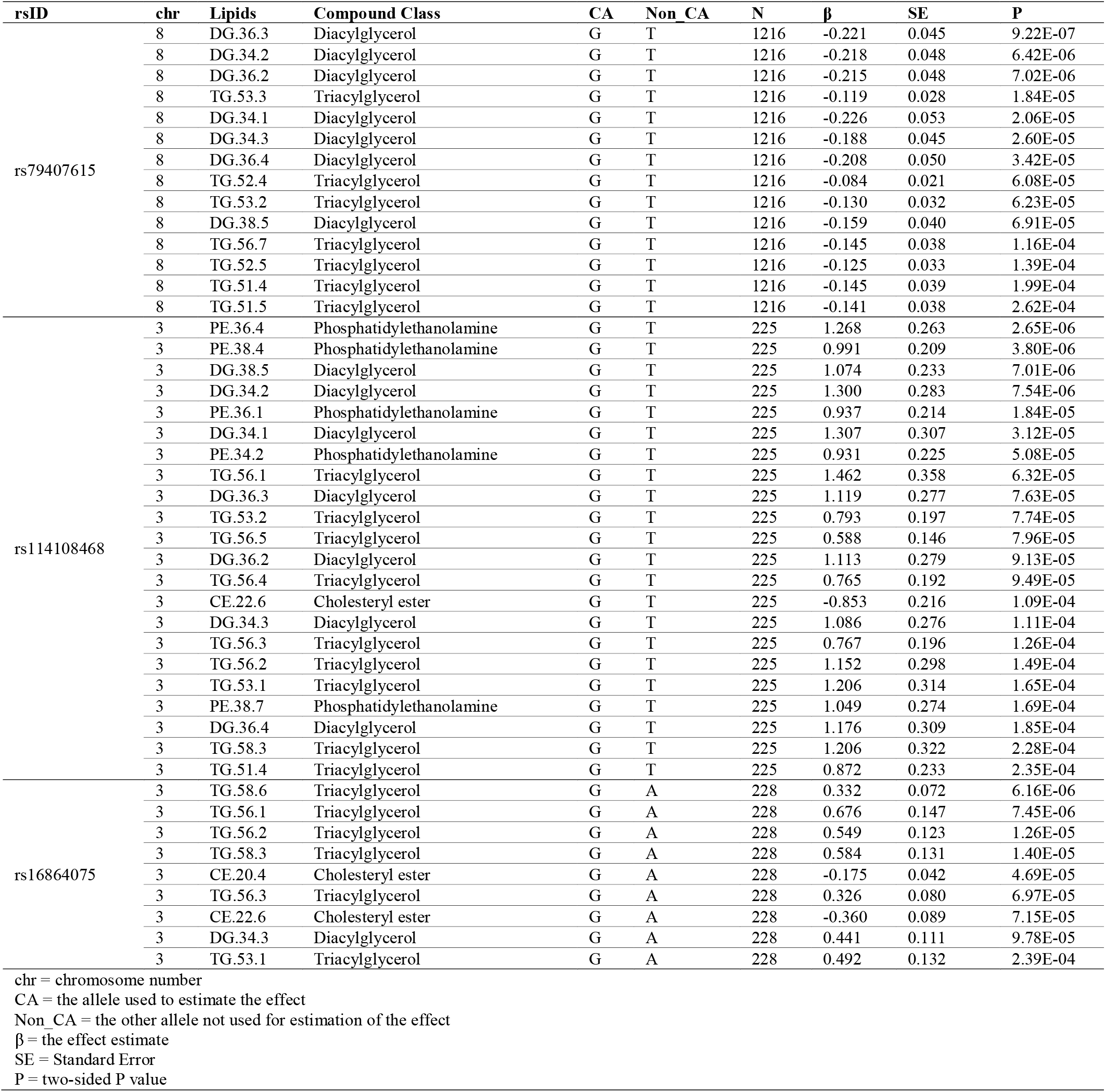
Lipids associated with LPL locus and 3q28 locus.

## Discussion

In our study, we characterized the genetic architecture underlying ΔTHR via two orthogonal approaches, GWAS and GWLS. In GWAS, we used a linear mixed model to account for the relatedness of 1384 LLFS participants and identified a genome-wide significant common variant of rs79407615 at chromosome 8 whose G allele is enriched in LLFS and associated with lower ΔTHR and THR. Within LLFS, this SNP is an eQTL for *LPL*, displaying a significant decreased blood *LPL* and 14 blood lipid level with the G allele. This enriched protective G allele might be the mechanism underpinning the lower incidence of T2D in LLFS. In line with our findings, *LPL* locus (rs7015766, rs254, rs268, rs276, rs73667496, and rs55682243) has been reported regulating cross-sectional THR [4]. Even though *LPL* locus (rs254, rs268, and rs276) has been reported for association with cross-sectional THR [4], the association of *LPL* locus with longitudinal ΔTHR in a healthy family cohort is novel and has not been identified before. In GWLS, we conducted oligogenic linkage analysis to model the association of shared genetic components in LLFS families with the similarity of ΔTHR. We found a significant novel linkage peak at 3q28 with LODs score exceeding 3. With 25 linkage enriched families (pedigree-specific LODs>0.1), we leveraged sequence data, and detected two novel variants (*EIF4A2*/*ADIPOQ*-rs114108468, *TPRG1*-rs168640750, each explaining approximately 28%-∼29% of the linkage. Notably, variant rs114108468 is an eQTL of *EIF4A2* and *ADIPOQ* in blood, indicating their involvement in regulating ΔTHR.

Our findings are supported by multiple lines of evidence. The *ADIPOQ* gene is involved in obesity, T2D, and body weight [20], with insulin-sensitizing and anti-atherogenic effects. The lower levels of *ADIPOQ* in blood observed in G allele carriers of rs114108468 might be one of the molecular mechanisms underlying the accelerated increase of THR of these individuals. *EIF4A2* (Eukaryotic Translation Initiation Factor 4A2) is involved in negative regulation of RNA-directed 5’-3’ RNA polymerase activity, and glucose homeostasis control. It has also been suggested as a potential candidate gene for T2D [21].

The association of this locus with other IR related traits are in line with their regulation in ΔTHR. For instance, multiple lipids were significantly associated with 3q28 locus. Several previous studies found suggestive evidence of linkage at 3q28 locus for adiponectin [22–25], dementia [26], Alzheimer’s disease [27], and Systolic Blood Pressure (SBP) [28], highlighting the pleiotropic roles of this locus for multiple traits.

To replicate the association of rs114108468 and rs16864075 with ΔTHR, we used the 859 FHS OS samples from offspring examinations 5 and 7. Even though these two SNPs did not reach nominal significance in FHS OS, we noted two nearby rare SNPs (rs189908673:11,998 bp downstream of rs114108468; rs113022387: 835 bp upstream of rs16864075) that are nominally significant in association with ΔTHR. This indicates that there are distinct variants within the identified locus regulating ΔTHR for FHS OS participants. The genetic and phenotypic difference among LLFS and FHS OS might be the reason that identification of different SNPs within the locus, which have a modest effect on regulating ΔTHR. LLFS samples were selected from exceptionally long-lived families, representing the upper 1% tail of the Family Longevity Selection Score across four field centers. In contrast, all FHS OS participants were residents of the city of Framingham, Massachusetts. Compared to FHS OS, LLFS participants exhibited relatively higher levels of HDL-c and lower levels of TG across all age categories (<60, 60-80, 80-100, and >100) [5]. Standardized ΔTHR was utilized in our analyses to address the disparity between LLFS and FHS OS. Given the differences in participant selection, it is likely that distinct genetic architectures underlie ΔTHR.

Despite these findings, it is important to acknowledge several limitations in our study. Firstly, owing to the relatively low allele frequency and the constraints of small sample size, our current analysis did not reveal molecules directly linked to rs16864075. Addressing these limitations would require future investigations, leveraging an expanded LLFS offspring generation and integrating multi-layer omic data, which will be instrumental in gaining a more comprehensive understanding of these genetic associations. Secondly, all LLFS samples are European descent, limiting the generalizability of our findings to other ancestries. Validating our results using samples from ancestries beyond European descent will be crucial to address this limitation. Thirdly, we cannot definitely conclude that the identified genetic variants are causal for ΔTHR without further functional experimental validation. Nonetheless, our analyses possessed several strengths. We utilized a family design, employed two orthogonal approaches, and integrated multi-omics data (genomic, transcriptomic and lipidomic).

In summary, by focusing on the genetic region under the linkage peak and utilizing the families selected for linkage, we uncovered novel variants and nominated potential genes (*EIF4A2, ADIPOQ, TPRG1*) influencing ΔTHR. The findings add new information in better understanding of pathophysiology of IR-associated chronic diseases and processes including T2D, AD and aging.

## Abbreviations

TG: Triglyceride
HDL-C: High-density lipoprotein cholesterol
THR: TG/HDL-C ratio
ΔTHR: TG/HDL-C ratio change over time
FHS: Framingham Heart Study
OS: Offspring Cohort
PCs: Principal components
IR: Insulin resistance
T2D: Type 2 Diabetes
eQTLs: Expression quantative trait loci
mQTLs: Metabolites quantative trait loci
LODs: Logarithm of the odds
HLOD: heterogeneity LOD

## Ethics declarations

## Data Availability

All data can be located at https://www.ncbi.nlm.nih.gov/projects/gap/cgi-bin/study.cgi?study_id=phs000397.v1.p1

## Acknowledgements

The authors would like to thank all the LLFS and FHS OS participants who agreed to contribute for scientific research. The authors also thank LeAnne Kniepkamp for her administrative contribution.

## Authors’ relationships and activities

The authors declare that there are no relationships or activities that might bias, or be perceived to bias, their work.

## Contribution statement

LW analyzed and drafted the manuscript. SW, JAA, VAM, and YL contributed to the analyses and provided edits. PA, JAA, ALK, MB, and BT revised the manuscript critically. SL contributed to quality control of the LLFS samples. PA and MP contributed to the conception and design of this manuscript. BT quantified the TG and HDL-C for LLFS. MB supervised the generation and quality control of RNA-seq data. GP was involved in the generation and quality control of blood lipid metabolite data. EWD, IM, and JMZ contributed to the LLFS data collection. All authors approved the final version of the manuscript.

